# Symptoms of COVID-19 infection and magnitude of antibody response in a large community-based study

**DOI:** 10.1101/2021.02.04.21251170

**Authors:** Thomas W. McDade, Joshua M. Schrock, Richard D’Aquila, Brian Mustanski, Nanette Benbow, Lauren A. Vaught, Nina L. Reiser, Matt E. Velez, Ryan R. Hsieh, Daniel T. Ryan, Rana Saber, Elizabeth M. McNally, Alexis R. Demonbreun

## Abstract

**Background:** The majority of COVID-19 cases are asymptomatic, or minimally symptomatic with management in the home. Little is known about the frequency of specific symptoms in the general population, and how symptoms predict the magnitude of antibody response to SARS-CoV-2 infection.

**Methods:** We quantified IgG antibodies against the SARS-CoV-2 receptor binding domain (RBD) in home-collected dried blood spot samples from 3,365 adults participating in a community-based seroprevalence study in the city of Chicago, USA, collected between June 24 and November 11, 2020.

**Results:** 17.8% of the sample was seropositive for SARS-CoV-2. A cluster of symptoms (loss of sense of smell or taste, fever, shortness of breath, muscle or body aches, cough, fatigue, diarrhea, headache) was associated with stronger anti-RBD IgG responses among the seropositives. 39.2% of infections were asymptomatic, and 2 or fewer symptoms were reported for 66.7% of infections. Total number of symptoms was positively but weakly associated with IgG response: Median anti-RBD IgG was 0.95 ug/mL for individuals with 3 or more symptoms, in comparison with 0.61 ug/mL for asymptomatic infections.

**Conclusion:** We document high rates of asymptomatic and mild infection in a large community-based cohort, and relatively low levels of anti-SARS-CoV-2 IgG antibody in the general population of previously exposed individuals.

## Introduction

A consequential feature of severe acute respiratory syndrome coronavirus 2 (SARS-CoV-2)—the virus that causes coronavirus disease 2019 (COVID-19)—is that a high proportion of infections are mild or asymptomatic.(1) The antibody response to COVID-19 is well characterized, and serological testing can be used to document prior infection by detecting the presence of antibodies against SARS-CoV-2 in blood samples.(2) Furthermore, the magnitude of antibody response correlates positively with the effectiveness of neutralization activity in virus challenge tests.(3) Therefore, quantitative assays for SARS-CoV-2 antibodies can be used to detect prior exposure—even in the absence of symptoms or a clinical diagnosis—and may provide information on the level of protection against future re-infection.

Relatively little is known about the frequency of COVID-19 symptoms in the general population, and how symptoms of SARS-CoV-2 infection relate to the magnitude of antibody response. Using data from a large community-based cohort in Chicago, we analyzed patterns of symptoms and their association with the magnitude of antibody response using a highly sensitive and quantitative assay for IgG antibodies against the receptor binding domain (RBD) of SARS-CoV-2. While important insights are gained from clinic-based studies of COVID-19, the majority of cases are asymptomatic, or minimally symptomatic and requiring only home-based treatment. A community-based approach allows us to estimate the prevalence of specific symptoms and their contribution to the development of immunity in the majority of infected persons who do not require hospitalization. We document high rates of asymptomatic and mild infection, relatively low levels of antibody response, and a dose-response relationship between flu-like symptoms and the magnitude of response.

## Methods

### Study design

The study uses data from Screening for Coronavirus Antibodies in Neighborhoods (SCAN), a community-based serological study in Chicago. Chicago residents were initially recruited from 10 zip codes geographically dispersed across the city, with enrollment opened city-wide in early September. Participants were also enrolled through the Feinberg School of Medicine in downtown Chicago. Participants were recruited through social media, email blasts from community organizations, print flyers, newspaper advertisements, and press coverage of the study in local media. A web-based, “no contact” research platform asked all participants to fill out a short questionnaire to determine study eligibility, followed by electronic informed consent and a longer survey. Upon completing the survey, a kit for collecting a finger stick dried blood spot (DBS) sample was mailed to the home or picked up at a designated time for onsite medical school participants. Instructions for self-collection were provided in print, and in an online video, and samples were mailed to the lab for analysis. All samples were de-identified and all research activities were implemented under protocols approved by the institutional review board at Northwestern University (#STU00212457 and #STU00212472).

### Measurement of SARS-CoV-2 IgG antibodies

Samples were analyzed for anti-RBD SARS-CoV-2 IgG antibodies with a quantitative enzyme-linked immunosorbent assay (ELISA) protocol that was previously validated for use with DBS.(4) The assay is based on a widely used protocol that has received Emergency Use Authorization from the US FDA,(5) and analysis of matched DBS and serum samples indicates near perfect agreement in results across the assay range.(4) The protocol uses known concentrations of a cross-reacting monoclonal antibody (CR3022) against RBD to generate a calibration curve from which values are calculated for unknowns. Based on prior validation, the cut-off for defining a sample as seropositive was set at 0.4 µg/mL or higher.

### Data analysis

The sample included N=3,365 participants who provided a DBS sample between June 24 and November 11, 2020. Participants were asked to report whether they experienced the following symptoms of infectious disease potentially indicative of COVID-19 after March 1, 2020: fever or chills; cough; shortness of breath; sore throat; headache; muscle or body aches; runny nose; fatigue or excessive sleepiness; diarrhea, nausea, or vomiting; loss of sense of smell or taste; itchy/red eyes. Other variables included pre-existing medical conditions (chronic obstructive pulmonary disease, diabetes mellitus, cardiovascular disease, obesity (body mass index >30 kg/m^2^,(6) smoking, and essential worker status (working outside the home in close proximity to others since March 1). Participants also indicated sex (based on assignment at birth), and racial/ethnic identity.

Descriptive statistics and multivariate regression analyses were used to identify patterns of association with IgG serostatus, and magnitude of IgG response to infection. Due to skew in the distribution of IgG values, median values are presented and log10 values were used in regression analyses. Polychoric correlation—which is appropriate for dichotomous ordinal variables—was used to analyze strength of association between individual symptoms and log IgG response, and to construct a correlation matrix for factor analysis of patterns of association among symptoms.(7) All analyses were implemented using Stata/SE, version 15.1 (StataCorp, College Station, TX). Since SCAN initially used a neighborhood-based study design which focused on 10 zip codes in Chicago, not all observations are independent and the cluster option in Stata was used to generate robust standard error estimates for multiple regression models, with zip code specified as the clustering variable.

## Results

Mean age of participants was 38.4 years (range: 18-86). 17.8% of the sample was seropositive for prior exposure to SARS-CoV-2. Seronegative and seropositive individuals reported the same overall frequency of symptoms. For both groups, the median number of symptoms was 2; however, the distributions were not equivalent, with a small upward shift for seropositive individuals (Wilcoxon rank-sum z=−2.64, p<0.01). Seropositive individuals were significantly more likely to report headache, fatigue, muscle or body aches, fever, and loss of sense of smell or taste (Table 1).

**Table 1.**
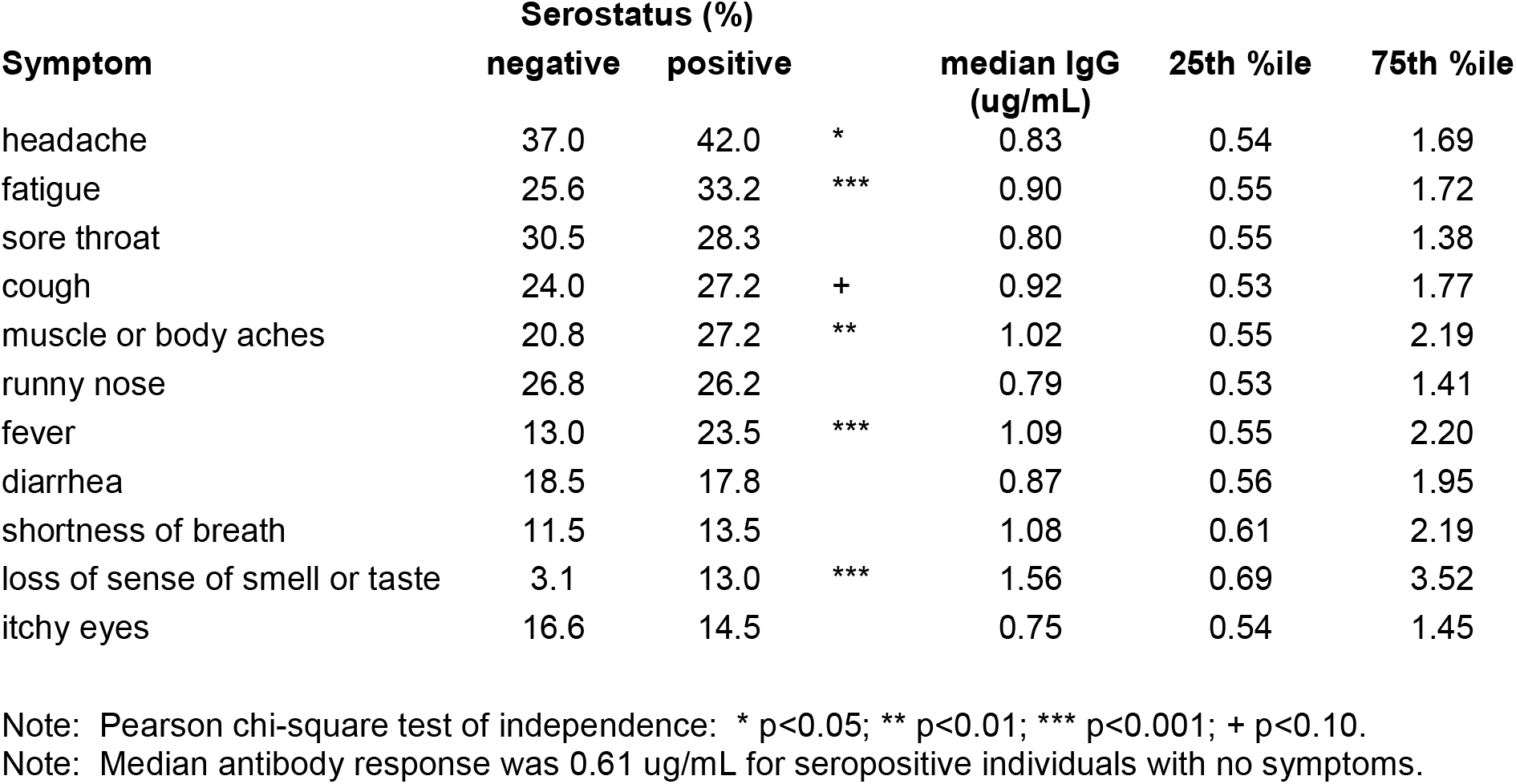
Reported symptoms of SARS-CoV-2 infection in association with seropositivity status and magnitude of anti-RBD IgG response for seropositive individuals.

| Symptom | Serostatus (%) |  |  | median IgG<br>(ug/mL) | 25th %ile | 75th %ile |
| --- | --- | --- | --- | --- | --- | --- |
|  | negative | positive |  |  |  |  |
| headache | 37.0 | 42.0 | * | 0.83 | 0.54 | 1.69 |
| fatigue | 25.6 | 33.2 | *** | 0.90 | 0.55 | 1.72 |
| sore throat | 30.5 | 28.3 |  | 0.80 | 0.55 | 1.38 |
| cough | 24.0 | 27.2 | + | 0.92 | 0.53 | 1.77 |
| muscle or body aches | 20.8 | 27.2 | ** | 1.02 | 0.55 | 2.19 |
| runny nose | 26.8 | 26.2 |  | 0.79 | 0.53 | 1.41 |
| fever | 13.0 | 23.5 | *** | 1.09 | 0.55 | 2.20 |
| diarrhea | 18.5 | 17.8 |  | 0.87 | 0.56 | 1.95 |
| shortness of breath | 11.5 | 13.5 |  | 1.08 | 0.61 | 2.19 |
| loss of sense of smell or taste | 3.1 | 13.0 | *** | 1.56 | 0.69 | 3.52 |
| itchy eyes | 16.6 | 14.5 |  | 0.75 | 0.54 | 1.45 |

Note: Pearson chi-square test of independence: \* $p < 0.05$ ; \*\* $p < 0.01$ ; \*\*\* $p < 0.001$ ; + $p < 0.10$ .
Note: Median antibody response was 0.61 ug/mL for seropositive individuals with no symptoms.

Among seropositive participants, median anti-RBD IgG concentration was 0.73 ug/mL (25^th^ %ile = 0.50, 75^th^ %ile = 1.32). For individuals reporting no symptoms, median IgG concentration was 0.61 ug/mL (25^th^ %ile = 0.47, 75^th^ %ile = 1.00). Loss of sense of smell or taste, fever, shortness of breath, muscle or body aches, cough, and fatigue were associated with the largest antibody response to infection, with median increases that were 1.5 to 2.6 times greater (Table 1). Sore throat, runny nose, and itchy eyes were associated with the smallest increases in antibody concentration.

Since infectious symptoms typically do not occur in isolation, we used factor analysis to identify patterns of clustering among symptoms. The first factor explains a high proportion of variance (0.874), with heaviest loadings for fatigue, muscle or body aches, fever, cough, and headache (Table S1). A second factor accounted for a smaller proportion of variance (0.137), and was comprised of three symptoms: runny nose, sore throat, and itchy eyes. Based on these results, we created a “cold-like symptoms” variable that summarized the presence of these three symptoms. We also created a “flu-like symptoms” variable that summarized all other symptoms.

Among seropositive individuals, 39.2% reported no flu-like symptoms of infection, and 34.7% reported neither flu-nor cold-like symptoms. Flu-like symptoms were a strong predictor of increased antibody response, particularly in the presence of three or more symptoms (Figure 1; Table S2). Age, sex, race/ethnicity, pre-existing conditions, smoking, and essential worker status were not significant predictors of the magnitude of antibody response, and their inclusion in the model did not reduce the association between flu-like symptoms and IgG concentration (Table S2).

**Figure 1.**
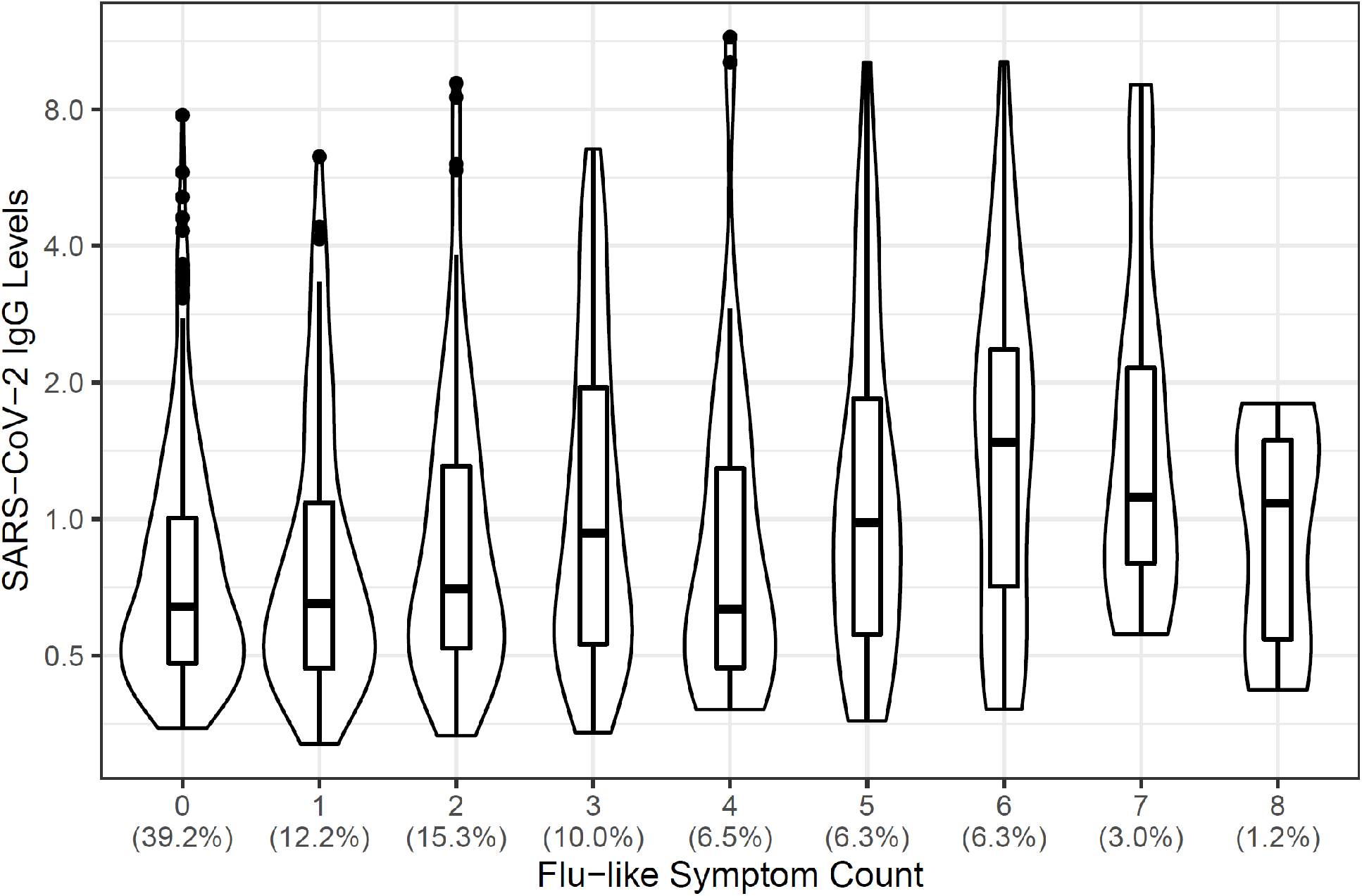
SARS-CoV-2 anti-RBD IgG concentration in association with number of flu-like symptoms. Symptoms include fever or chills, cough, shortness of breath, headache, muscle or body aches, fatigue or excessive sleepiness, diarrhea, nausea, or vomiting; loss of sense of smell or taste. Violin plot shows median IgG and interquartile range, with kernel density, at each symptom level. Proportion of seropositive individuals at each symptom level is presented below the x-axis. The y-axis is presented on a binary logarithmic scale.

The dose-response association between flu-like symptoms and anti-RBD IgG antibody response is positive but relatively weak. Further, many asymptomatic infections produced strong antibody responses, and conversely, many symptomatic infections resulted in relatively weak antibody responses. The median response for participants with 3 or more flu-like symptoms was 0.95 ug/mL, in comparison with 0.61 ug/mL for those with no symptoms. However, 33.0% of participants with 3 or more flu-like symptoms had IgG<0.61 ug/mL, while 27.2% of asymptomatic infections produced IgG>0.95 ug/mL.

## Discussion

Symptoms potentially indicative of COVID-19 infection were recognized early in the pandemic, and symptom reports have been used to identify probable cases, to prioritize access to diagnostic testing, and to mobilize preventive behaviors like self-isolation and mask wearing. In a large, community-based serosurvey, we find comparable levels of overall symptom reporting among seropositive and seronegative individuals since March 1, 2020. However, a subset of flu-like symptoms were significantly associated with the likelihood of infection, and with the magnitude of antibody response following infection.

Most testing for SARS-CoV-2 virus has targeted those with symptoms or recent exposure. In the absence of community-based screening programs, serological testing is an important tool for identifying the correlates of SARS-CoV-2 infection in the community. In the dense urban context of Chicago, we find a high prevalence of seroconversion to SARS-CoV-2 at 17.8 percent, and a rate of asymptomatic infection near 40 percent among those with detectable antibody. Two or fewer symptoms were reported for two-thirds of all seropositives.

More flu-like symptoms predict higher antibody responses, but the magnitude of response is small in this community-based sample. By comparison, using the same DBS assay applied in this study, we have reported a median anti-RBD IgG response of 98.5 ug/mL for patients hospitalized with confirmed COVID-19 (8)—more than 10 times greater than the median response for individuals with 3 or more flu-like symptoms. To the extent that anti-RBD IgG concentrations are positive predictors of neutralizing activity against SARS-CoV-2,(3) our findings suggest a relatively low level of immunity in the general population of previously exposed individuals, the vast majority of whom experience no or minimal symptoms and do not require hospitalization.

## Data Availability

Data are available upon request from the corresponding author subject to an approved data sharing agreement to protect participant confidentiality.

---

**Table S1.** Factor loadings for symptoms of SARS-CoV-2 infection.

| Symptom | Factor 1 | Factor 2 | Uniqueness |
| --- | --- | --- | --- |
| fever | 0.816 | -0.297 | 0.245 |
| cough | 0.806 | 0.064 | 0.346 |
| shortness of breath | 0.683 | -0.101 | 0.523 |
| sore throat | 0.578 | 0.399 | 0.507 |
| headache | 0.800 | 0.086 | 0.353 |
| muscle or body aches | 0.840 | -0.315 | 0.196 |
| runny nose | 0.559 | 0.457 | 0.479 |
| fatigue | 0.846 | -0.006 | 0.285 |
| diarrhea | 0.665 | -0.070 | 0.553 |
| loss of smell or taste | 0.657 | -0.293 | 0.483 |
| itchy eyes | 0.465 | 0.458 | 0.574 |

**Table S2.** Ordinary least squares regression results for the association between symptoms of SARS-CoV-2 infection and log10 anti-RBD IgG concentration among seropositive individuals (n=600).

| Variable | B | 95% CI |  |  | B | 95% CI |  |  | B | 95% CI |  |  |
| --- | --- | --- | --- | --- | --- | --- | --- | --- | --- | --- | --- | --- |
| Flu-like symptoms | 0.036 | 0.024 | 0.047 | ** |  |  |  |  | 0.041 | 0.027 | 0.056 | ** |
| Cold-like symptoms |  |  |  |  | 0.016 | -0.014 | 0.046 |  | -0.041 | -0.072 | -0.009 | * |
| Age group<sup>1</sup>: <25 years |  |  |  |  |  |  |  |  | 0.017 | -0.051 | 0.085 |  |
| 35 to <45 |  |  |  |  |  |  |  |  | -0.020 | -0.083 | 0.043 |  |
| 45 to <55 |  |  |  |  |  |  |  |  | 0.063 | -0.023 | 0.148 |  |
| 55 to <65 |  |  |  |  |  |  |  |  | -0.023 | -0.126 | 0.080 |  |
| 65 and over |  |  |  |  |  |  |  |  | -0.046 | -0.183 | 0.092 |  |
| Sex |  |  |  |  |  |  |  |  | 0.002 | -0.046 | 0.050 |  |
| Race/ethnicity<sup>2</sup>: Asian |  |  |  |  |  |  |  |  | -0.051 | -0.114 | 0.012 |  |
| Black |  |  |  |  |  |  |  |  | 0.047 | -0.053 | 0.147 |  |
| Hispanic |  |  |  |  |  |  |  |  | 0.010 | -0.055 | 0.075 |  |
| Other |  |  |  |  |  |  |  |  | 0.129 | -0.046 | 0.304 |  |
| Pre-existing conditions |  |  |  |  |  |  |  |  | -0.001 | -0.045 | 0.043 |  |
| Current smoker |  |  |  |  |  |  |  |  | 0.005 | -0.088 | 0.097 |  |
| Essential worker |  |  |  |  |  |  |  |  | -0.005 | -0.058 | 0.047 |  |
| Constant | -0.120 | -0.150 | -0.090 | ** | -0.060 | -0.100 | -0.030 | ** | -0.104 | -0.197 | -0.011 | * |

<sup>1</sup>The omitted group is age 25 to <35<sup>2</sup>The omitted group is "white"
\*\*p-value<0.01; \*p-value<0.05

## Notes

### Competing Interest Statement

Dr. McDade reports that he has a financial interest in EnMed Microanalytics, a company that conducts lab tests using DBS samples.

Dr. D’Aquila reports personal fees from Abbvie, outside the submitted work.

Dr. McNally reports personal fees from Amgen, personal fees from AstraZeneca, personal fees from Cytokinetics, personal fees from Pfizer, personal fees from Tenaya Therapeutics, personal fees from 4D Molecular Therapeutics, outside the submitted work.

### Funding Statement

This material is based upon work supported by the National Science Foundation under Grant No. 2035114. Additional support was provided by the Northwestern University Office of Research, the Northwestern University Clinical and Translational Sciences Institute (NIH UL1TR001422), and a generous gift from Dr. Andrew Senyei and Noni Senyei. The funding sources had no role in the study design, data collection, analysis, interpretation, or writing of the report.

### Author Declarations

All samples were de-identified and all research activities were implemented under protocols approved by the institutional review board at Northwestern University (#STU00212457 and #STU00212472).

## References

1. Oran DP, Topol EJ. Prevalence of asymptomatic SARS-CoV-2 infection : A narrative review. Ann Intern Med. 2020;173(5):362–7.

2. World Health Organization. Population-based age-stratified seroepidemiological investigation protocol for COVID-19 virus infection, 17 March 2020. World Health Organization; 2020.

3. Wajnberg A, Amanat F, Firpo A, Altman DR, Bailey MJ, Mansour M, et al. Robust neutralizing antibodies to SARS-CoV-2 infection persist for months. Science. 2020;370(6521):1227–30.

4. McDade T, McNally E, Zelikovich A, D’Aquila R, Mustanski B, Miller A, et al. High seroprevalence for SARS-CoV-2 among household members of essential workers detected using a dried blood spot assay. PLoS One. 2020;15(8):e0237833.

5. Amanat F, Nguyen T, Chromikova V, Strohmeier S, Stadlbauer D, Javier A, et al. A serological assay to detect SARS-CoV-2 seroconversion in humans. MedRxiv. 2020. https://doi.org/10.1101/2020.03.17.20037713

6. Centers for Disease Control and Prevention. COVID-19: People with Certain Medical Conditions 2021 [Available from: https://www.cdc.gov/coronavirus/2019-ncov/need-extra-precautions/people-with-medical-conditions.html.

7. Kolenikov S, Angeles G. The use of discrete data in PCA: theory, simulations, and applications to socioeconomic indices. Chapel Hill: Carolina Population Center, University of North Carolina. 2004;20:1–59.

8. Demonbreun AR, McDade TW, Pesce L, Vaught LA, Reiser NL, Bogdanovic E, et al. Patterns and persistence of SARS-CoV-2 IgG antibodies in a US metropolitan site. medRxiv. 2020. https://doi.org/10.1101/2020.11.17.20233452

